# Polygenic scores capture genetic modification of the adiposity-cardiometabolic risk factor relationship

**DOI:** 10.1101/2025.04.09.25324066

**Authors:** Kenneth E. Westerman, Julie E. Gervis, Luke J. O’Connor, Miriam S. Udler, Alisa K. Manning

## Abstract

Optimal use of genetics for precision medicine requires polygenic scores (PGS) that predict not just risk of disease, but also response to pharmaceutical or lifestyle interventions. These are detectable in observational datasets as PGS-by-exposure (PGS×E) interactions. Existing literature suggests that PGS based on interactions (iPGS) or variance effects (vPGS) may be more powerful than standard marginal PGS (mPGS) for the detection of PGS×E, but these have yet to be systematically compared. We describe a generalized pipeline for the development and comparison of different PGS types and apply it to detect genetic modification of the relationship between adiposity (measured by body mass index [BMI]) and a broad set of cardiometabolic risk factors (CRFs). Our applied analysis in the UK Biobank cohort identified significant PGS×BMI for at least one PGS type for 16/20 of these CRFs, many of which replicated in the All of Us cohort. Among PGS types, iPGS uncovered interactions with BMI most consistently across CRFs, with the strongest interactions impacting biomarkers of liver function (e.g., alanine aminotransferase [ALT]). Exploring the ALT iPGS more in-depth, we find a substantial effect modification of up to 72% larger BMI-ALT association in the top iPGS decile in All of Us, and further provide evidence that the iPGS prioritizes variants affecting hepatic lipid export. Taken together, our study provides a framework for the development and comparison of PGS×E strategies, quantifies genetic impacts on the adiposity-cardiometabolic risk relationship, and informs efforts to move toward clinically useful response-focused PGS.

## Introduction

Clinical decision-making is often based on risk estimates, in which patients at higher risk for a disease are prioritized for lifestyle changes or pharmaceutical treatments. However, individuals can vary widely in their response to these clinical interventions^1^, motivating the use of molecular measurements to predict therapeutic response and enable more targeted treatment recommendations. Genetic factors contribute to this inter-individual heterogeneity via gene-environment interactions (G×E), which quantify genetic effects on the association between some exposure (e.g., a behavior or pharmacological treatment) and the outcome of interest. As with standard risk prediction, G×E testing in epidemiological contexts can uncover stronger effects by combining information from variants across the genome using polygenic scores (PGS).

Several types of genome-wide statistical tests have been described for the development of PGS for G×E testing. Summary statistics from a genome-wide association study can be used to produce a standard marginal PGS (mPGS). This approach has successfully detected polygenic G×E^2–4^, but requires a strong and typically unsatisfied assumption that genetic main effects and interaction effects are proportional genome-wide^5^. Alternatively, G×E effects from a genome-wide interaction study can be used to produce an interaction PGS (iPGS). Prior studies have shown that iPGS can improve genetic prediction performance (explaining more outcome variability)^6–11^ and predict response to interventions^5,12^. Finally, genome-wide variance-quantitative trait locus (vQTL) analysis tests genetic associations with the variability, rather than the mean, of quantitative traits and can appear as the result of underlying interactions^13,14^. These summary statistics can be aggregated into variance PGS (vPGS)^15–17^, which have the advantage that their development does not require the explicit modeling of often poorly-measured exposure variables.

These PGS types have not yet been compared for their detection of interactions in a systematic way. Here, we propose a generalized pipeline for the generation of PGS for PGS×E testing and compare the performance of PGS based on each of these three genome-wide association models. We hypothesize that, by more directly quantifying effect modification, the iPGS and vPGS will detect stronger PGS×E interactions compared to the mPGS. We first explore this hypothesis through simulations, focusing on the impact of exposure distribution and measurement error on the relative performance of these PGS types for detecting interactions. We then conduct extensive analysis of genome-wide genetic modification of the strong known relationship between adiposity and cardiometabolic risk factors (CRFs) in the UK Biobank (UKB) and All of Us (AoU) datasets. As an exposure, we use body mass index (BMI), a measure of adiposity that strongly predicts chronic disease risk and participates in G×Es at the single-variant^18,19^ and PGS^11,20^ levels. As outcomes, we use a set of 20 continuous serum CRFs capturing a broad cross-section of physiological processes and genetic architectures^21^. The resulting PGS for each CRF will thus quantify the expected degree of change in that CRF in response to weight change.

## Results

### Conceptual overview of the PGS generation pipeline

The analysis pipeline depicted in Fig. 1 includes three steps: genome-wide association testing, PGS generation and optimization, and PGS×E testing. Genome-wide scans were performed using each of three statistical approaches. The first is the standard genome-wide association study (GWAS):

**Figure 1:**
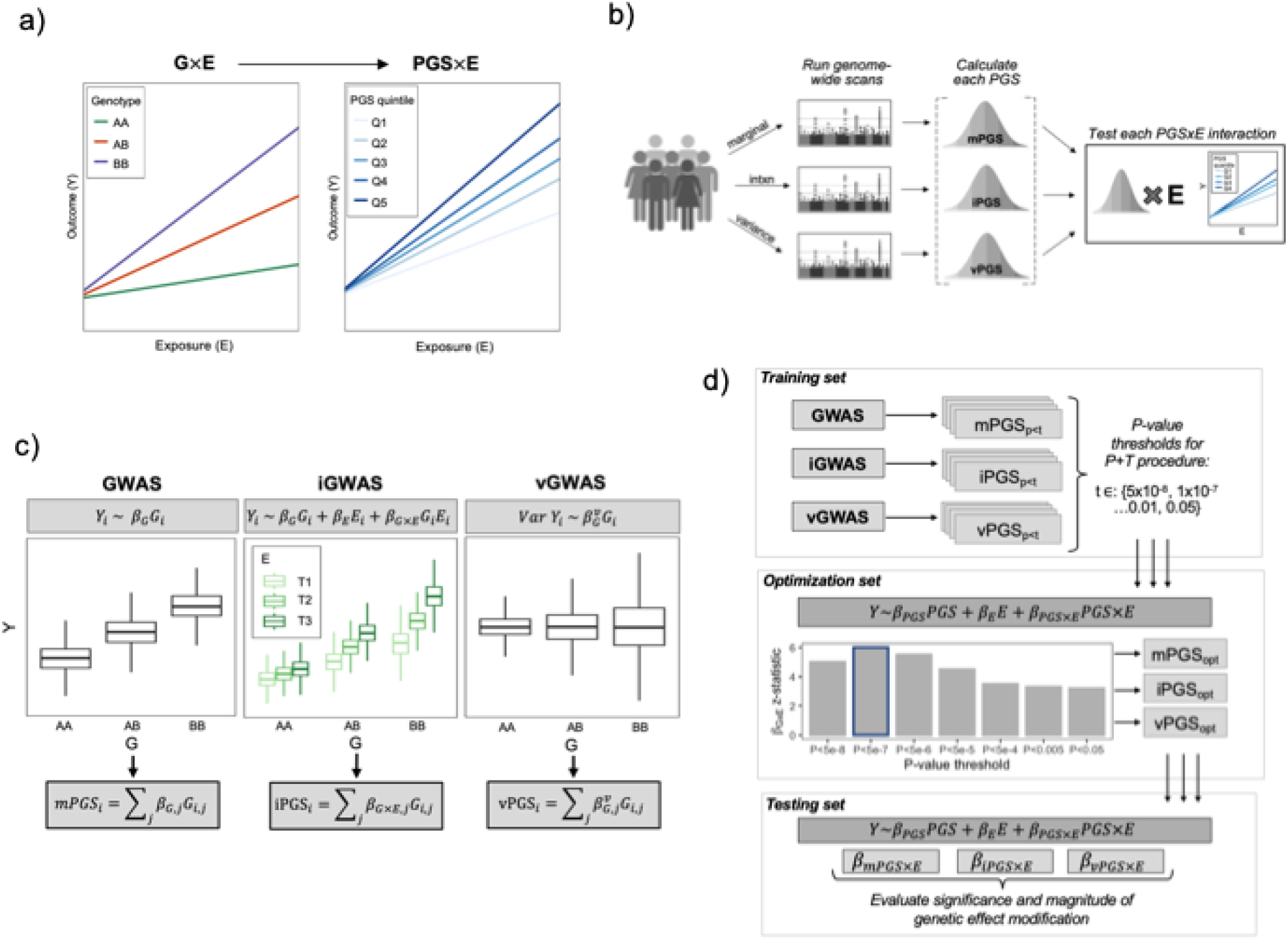
Conceptual overview of the analytical approach. (a) A PGS for response, developed using observational data, generalizes the concept of a G×E from a single genetic variant to a continuous, multi-variant score. (b) Various genome-wide scans, using individual-level data, might produce PGS that interact with a given exposure, with the optimal choice possibly depending on the specific biological question. (c) Multiple regression approaches can be used in the genome-wide scan, including genetic main effects (GWAS), interaction effects (iGWAS), and genetic variance effects, i.e., variance-quantitative loci (vGWAS). (d) Practical illustration of this pipeline for one outcome biomarker, using a P&T strategy. Genome-wide summary statistics are first generated in a training data subset and used to develop associated PGS at a series of *p*-value thresholds. These PGS are tested for interaction with the exposure in the optimization subset to select an optimal threshold for each PGS type based on the significance of the interaction effect, *β*_*PGSXE*_. Finally, these optimized PGS are each tested in a similar regression in the held-out testing subset and compared based on the same *β*_*PGSXE*_ estimate.

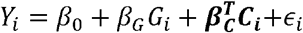

Where *Y*_*i*_ is the outcome for individual *i, G*_*i*_ is the genotype vector, *C*_*i*_ is a vector of covariates, and ϵ_*i*_ captures residual error. The genome-wide interaction study (iGWAS) model is a straightforward extension of the GWAS model:

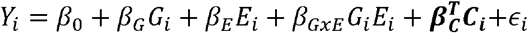

Where additional terms have been added for *E*_*i*_, the exposure, and its product term with *G*_*i*_. The key estimate of interest from this model is *β*_*GxE*_ (the interaction effect), rather than *β*_*G*_. Finally, the genome-wide vQTL study (vGWAS) models genetic effects on trait variability, which can capture underlying interactions without directly modeling the exposure. This produces an estimate of variability change per allele, denoted here as 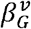 (see Methods).

For each set of genome-wide summary statistics (*β*_*G*_, *β*_*GxE*_, and 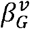), a pruning-and-thresholding (P&T) algorithm is used to develop a group of PGS based on different *p*-value thresholds. We note that more advanced PGS algorithms would be conceptually applicable, but will require additional methods and software development to optimize for the detection of interaction effects. An independent optimization data subset is then used to test for PGS×E interactions to select the optimal *p*-value threshold for each PGS type:

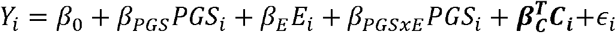

Where the optimal threshold is chosen as the one maximizing the significance of *β*_*PGSxE*_. Finally, a third independent testing data subset uses the same regression model to evaluate the significance and clinical significance of the *β*_*PGSxE*_ effect. This effect, estimated in an independent dataset, quantifies the expected change in the exposure-outcome relationship for each unit increase in the PGS and allows the direct comparison of the performance of the mPGS, iPGS, and vPGS strategies.

### Simulation results

Prior simulations studies exploring the use of mPGS and iPGS for detecting PGS×E interactions have reported several key results. Control of type I error for PGS×E testing requires adjustment for the main effect of the PGS of interest in the context of G-E correlation and residual-environment interaction (i.e., heteroscedasticity), as well as an additional permutation-based testing approach in some contexts^22^. Additionally, the relative power of the iPGS compared to the mPGS improves as the correlation between genetic main and interaction effects decreases^5^. Here, we sought to expand on a few specific components. First, we integrated the vPGS into type I error analyses within a single simulation and analysis pipeline. Second, we explored nonnegative exposure distributions. Though standard normal exposures are the default in simulations, they do not reflect most true biological quantities and generative models including negative exposure values can produce interactions that are difficult to interpret and potentially unrealistic^23^. Third, we added simulated exposure measurement error into power comparisons between the three approaches, acknowledging that this is a major challenge for GxE studies^24^. The simulation strategy is described in detail in the Methods and in Supp. Fig. S1.

In type I error analyses using a standard normally-distributed E, we found that type I error was controlled for all PGS types regardless of the presence of G or E main effects (Fig. 2a). Using a nonnegative (gamma-distributed) E, type 1 error remained controlled and was robust to G-E correlation (Fig. 2b). However, in keeping with theoretical expectations for single-variant G×E analysis, type I error became inflated when this G-E correlation was combined with a nonlinear E-Y relationship^25^ (Fig. 2c). Thus, these type I error findings largely recapitulated existing observations while validating the control of false positives using the analytical pipeline deployed in this study.

**Figure 2:**
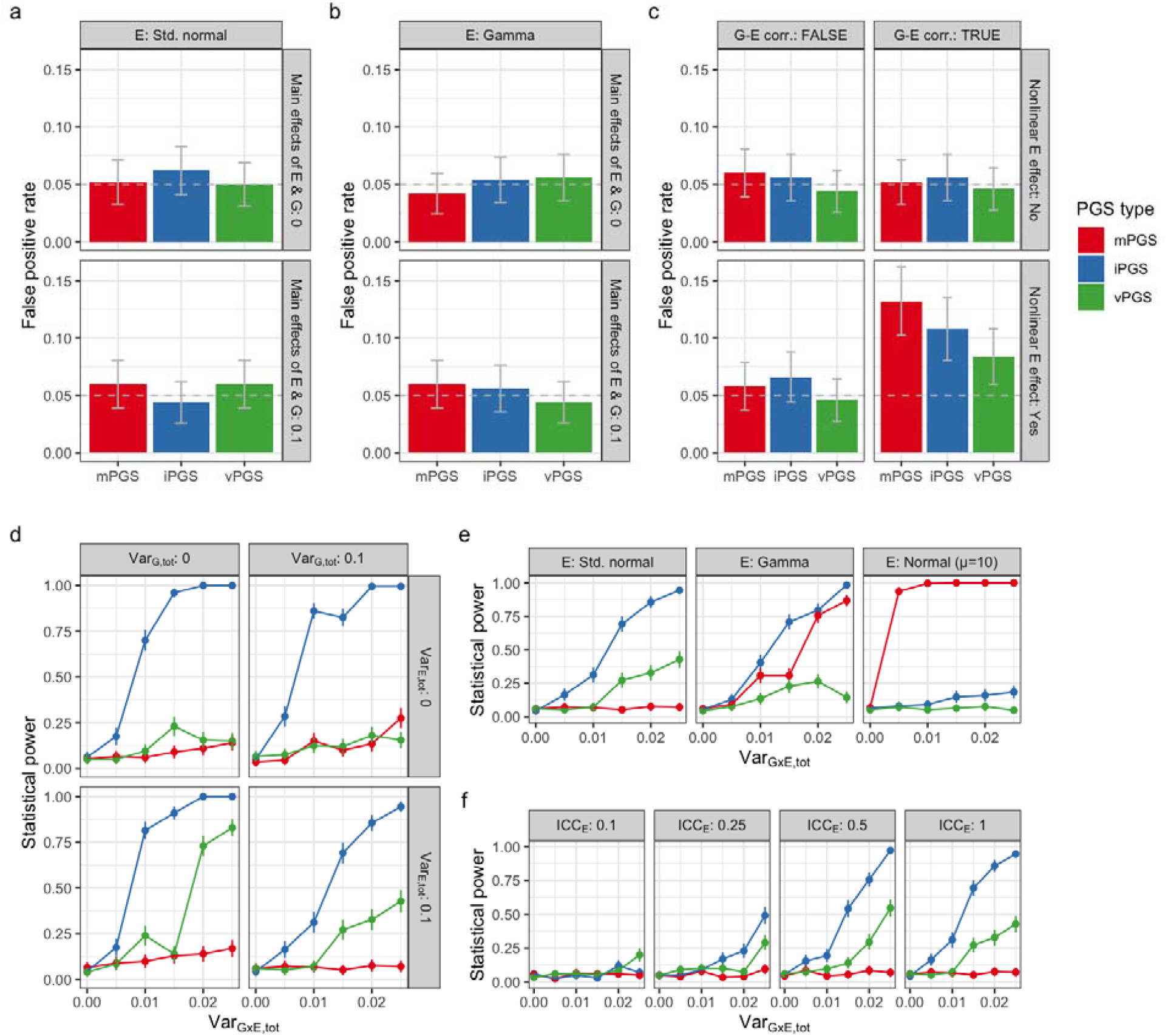
Simulation studies. (a-c) Type I error plots showing the false positive rate for the detection of PGS×E interaction when there is no underlying simulated G×E effect. Scenarios include a standard normally distributed E (a) and a gamma-distributed E (b). In the gamma-distributed case, the effect of G-E correlation and a nonlinear E-Y relationship were further modified (c). (d-f) Power plots show the performance of each PGS type in detecting PGS×E interaction as the variability due to G×E (*x*-axis) is manipulated. Scenarios include manipulation of G and E main effects with a standard normal E (d), manipulation of the E distribution (f), and manipulation of the simulated E measurement error with a standard normal E (e). ICC: intraclass correlation coefficient.

In power analyses using a standard normally-distributed E and assuming no correlation between simulated genetic main and interaction effects, we found that the iPGS approach was broadly the most powerful, followed by vPGS and finally mPGS (Fig. 2d). This matches expectations, given that the associated statistical test most closely matches the underlying simulated G×E interaction effects. When using E distributions with nonzero means, the mPGS became more relatively powerful: moderately so for a gamma-distributed E, and strongly so for a normally-distributed E with mean equal to 10 (Fig. 2e). As has been discussed in depth in the literature, an increasing mean of E raises the correlation of G and G×E product terms and thus the power of the marginal genetic test for detecting G×E^23^. Exposure measurement error hurt the statistical power of all approaches (Fig. 2f), but with less proportional impact on the vPGS (which detects interaction patterns without explicitly testing the exposure during PGS development).

### Primary PGS development and testing in the UK Biobank

The UKB dataset was used for the primary applied data analysis portion of this investigation. A summary of the relevant multi-ancestry, unrelated subgroup of the UKB population is provided in Supp. Table S1, including within the relevant training (70%), optimization (10%), and testing (20%) subsets. First, GWAS, iGWAS, and vGWAS were conducted for each CRF (see Methods). For each of these approaches, summary statistics were LD-pruned and PGS were generated corresponding to a series of *p*-value thresholds. This P&T strategy enabled PGS optimization (i.e., choice of optimal P&T *p*-value threshold) based on the strength of PGS×BMI interaction, rather than the PGS main effect (see Methods).

Next, a single optimal PGS was created for each combination of CRF and approach by choosing the *p*-value threshold optimizing the significance of the *β*_*PGSxBMI*_ regression term in the optimization subset (Supp. Fig. S2; all PGS weights are provided in the Supplemental Materials). Finally, the PGS performance was evaluated based on the magnitude and significance of the same *β*_*PGSxBMI*_ term in the testing subset, adjusting for covariates including basic demographics and genetic principal components. As a positive control, we confirmed that this data splitting and PGS development pipeline produced mPGS with strong marginal effects in the testing subset (Supp. Fig. S3).

Significance for the primary estimates was assigned based on a Bonferroni threshold adjusting for 10.3 effective CRFs as previously described for analysis of many biomarkers in the UKB (see Methods)^14,26^. Of 20 total CRFs, 16 passed the significance threshold for at least one PGS type. The iPGS reached Bonferroni significance for the greatest number of CRFs (Fig. 3b) and generated the most significant PGS×BMI interaction for 11 of the 16 CRFs that reached Bonferroni significance for any approach.

**Figure 3:**
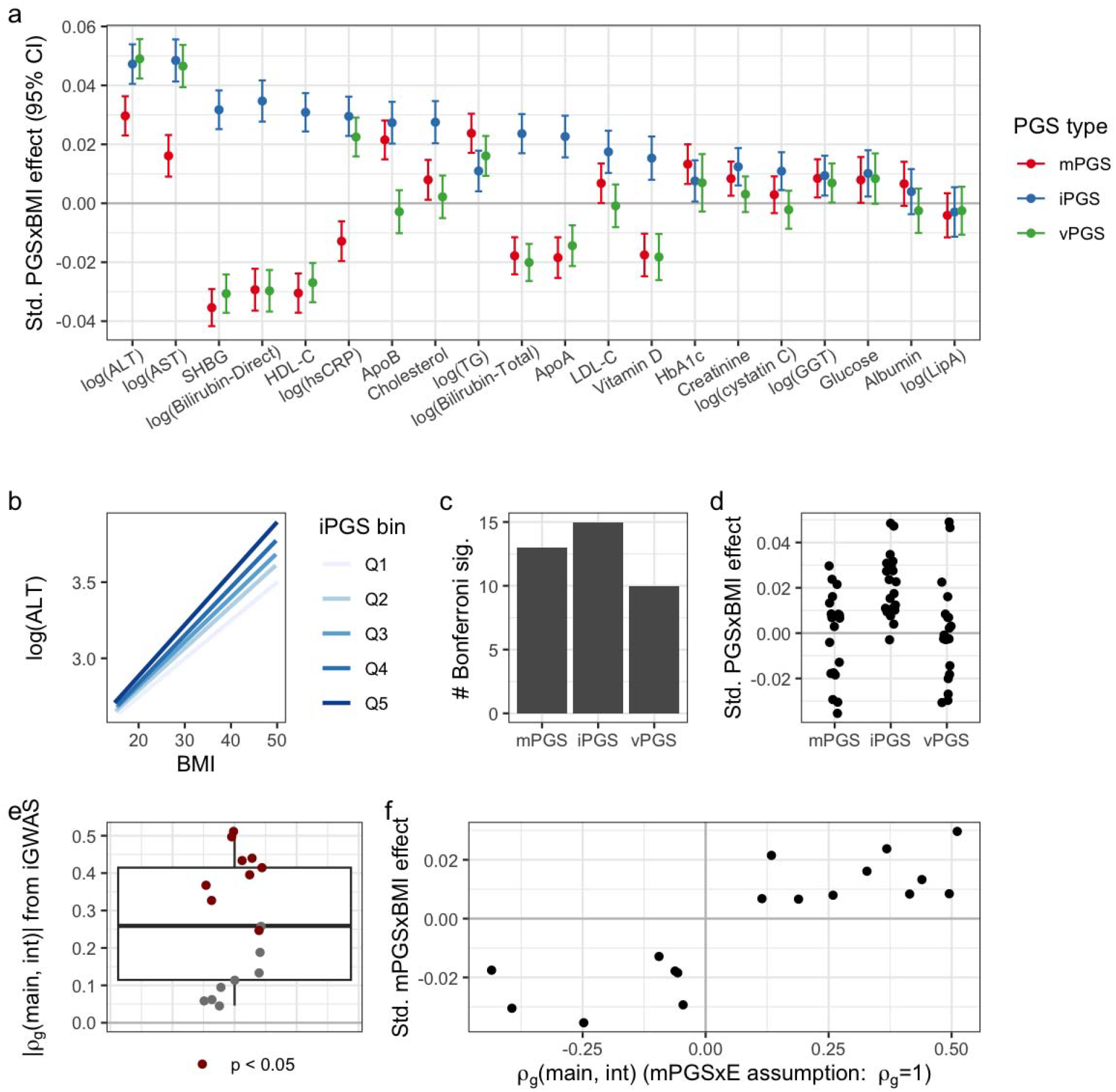
Primary results for optimized PGS in the UKB testing set. (a) Standardized interaction estimates (units of *SD*_*Y*_ /*SD*_*PGS*_ / *SD*_*BMI*_) are plotted for all CRFs (*x*-axis) and PGS types (colors). (b) Example stratified plot showing best fit lines for the relationship between BMI and log(ALT) within quintiles of the ALT iPGS. (c) Number of CRFs reaching Bonferroni significance for each approach. (d) Standardized interaction estimates for each approach. (e) Boxplot and individual points representing the absolute values of the genetic correlation between iGWAS main and interaction effects, as computed by LD-score regression. (f) Interaction effect for the mPGS is plotted against signed genetic correlation estimates.

Of the three PGS types, the iPGS approach most frequently captured the strongest BMI interactions across CRFs. This was not solely explained by better performance on a single, large cluster of correlated CRFs (see Supp. Fig. S4a). The mPGS approach showed substantial negative interaction estimates for some CRFs (i.e., higher mPGS leads to a decreased BMI-CRF association; Fig. 3c), notably for many having an inverse relationship with BMI (Supp. Fig. S4). This observation fits with a previously described pattern of genetic effect amplification by adverse exposures^4^ (see Discussion for further commentary). The vPGS showed negative interaction estimates for the same set of CRFs, consistent with the strong known relationship between genetic main and variance effects^15,26^. Though there were substantial vPGS×E interactions for some CRFs (e.g., ALT and AST), the vPGS did not meaningfully improve upon the iPGS for any of these.

As previously noted, the value of mPGS for the detection of G×E is directly related to the proportionality of genetic main and interaction effects. We quantified this directly by calculating genetic correlations (*ρ*_*g*_) between main and interaction effects from the same iGWAS using LD-score regression (Supp. Table S4). Of 17 CRFs with *ρ*_*g*_ estimates (interaction signal was insufficient for *ρ*_*g*_ estimation for three), the magnitude of these correlations ranged from 0.05 (nonsignificant; bilirubin direct) to 0.51 (*p* = 5.7×10^−14^) (Fig. 3e). These magnitudes are much smaller than the perfect correlation assumed by the mPGS approach, agreeing with results from Zhai and colleagues in a different domain of gene-statin interactions^5^. Though mPGS interaction effect signs matched *ρ*_*g*_ signs, there was no correlation between the magnitude of these quantities as might be expected theoretically (Pearson correlation between |*ρ*_*g*_|and |*β*_*mPGSXBMI*_| of 0.02; *p* = 0.9) (Fig. 3f).

### Sensitivity analyses in the UK Biobank

As demonstrated in our simulation study, a key concern in G×E testing is that interactions can appear as a statistical artifact of the combination of G-E correlation and nonlinearity of the E-Y relationship^25^. This issue is particularly relevant in this application given the highly polygenic nature of BMI. However, sensitivity models including either nonlinear BMI effects (squared BMI main effect term), nonlinear PGS effects, or using robust standard errors did not affect the results (Supp. Fig. S5a-c). Furthermore, when swapping out the iPGS in favor of an mPGS for BMI (which maximizes the achievable PGS-BMI correlation), the results were less strong for most CRFs (Supp. Fig. S5d). Together, these results suggest that PGS-BMI correlation is not solely responsible for the observed interaction effects.

We used further sensitivity models to explore two questions relevant to PGS×E testing. First, when testing iPGS for interaction, it may be valuable to adjust for the main effect of an mPGS in addition to the existing iPGS main effect. This could explain additional variability in the outcome and thus improve the significance of the interaction estimate (Jayasinghe and colleagues^22^; personal communication, D. Jayasinghe). We wanted to avoid this sort of adjustment for multiple PGS in our primary models for maximal interpretability, but we ran a set of sensitivity analyses including mPGS main effects in iPGS×BMI interaction models (in addition to the iPGS main effect already present in the model). This adjustment did not meaningfully affect the interaction results (Supp. Fig. S6).

Second, when using an exposure such as BMI that is under strong genetic influence, it is possible to replace the measured exposure with a PGS for that exposure before testing the interaction; ultimately, this results in a form of G×G test. This may be useful in two ways: it can strengthen the causal inference (by using a genetic causal anchor for BMI) and might reveal stronger underlying interactions occurring “upstream” of realized BMI (Supp. Fig. S7a,b). To test this, we tested each of the primary iPGS interaction models after replacing measured BMI with an mPGS for BMI (the same one used to replace the iPGS above). For most CRFs, the resulting iPGS×mPGS_BMI_ interactions were nonzero but less significant than those using measured BMI (Supp. Fig S7c). This finding is consistent with these interactions involving true causal effects of BMI that are not limited to its genetic component.

### Replication in All of Us

PGS were calculated in AoU based on optimized UKB variant weights for each approach-CRF combination (population summary in Supp. Table S5; biomarker metadata in Supp. Table S6). Regressions mirroring those in UKB were then run to understand how these scores generalize to a fully independent dataset and population (regression results in Supp. Table S7). We saw replication (at nominal *p* < 0.05) of many of the PGS interactions in the primary, pooled-ancestry dataset: 5/11 for mPGS, 6/13 for iPGS, and 3/8 for vPGS (Fig. 4a,c; full set of results comparing UKB and AoU in Supp. Table S8). PGS×BMI interaction effect sizes were strongly associated between the two cohorts, with Pearson correlations of 0.54, 0.70, and 0.69 for the mPGS, iPGS, and vPGS, respectively (Fig. 4b). Some of the higher-level patterns observed in UKB were also seen in AoU, including the general outperformance of the iPGS and a directionality of mPGS interaction effects consistent with the amplification model.

**Figure 4:**
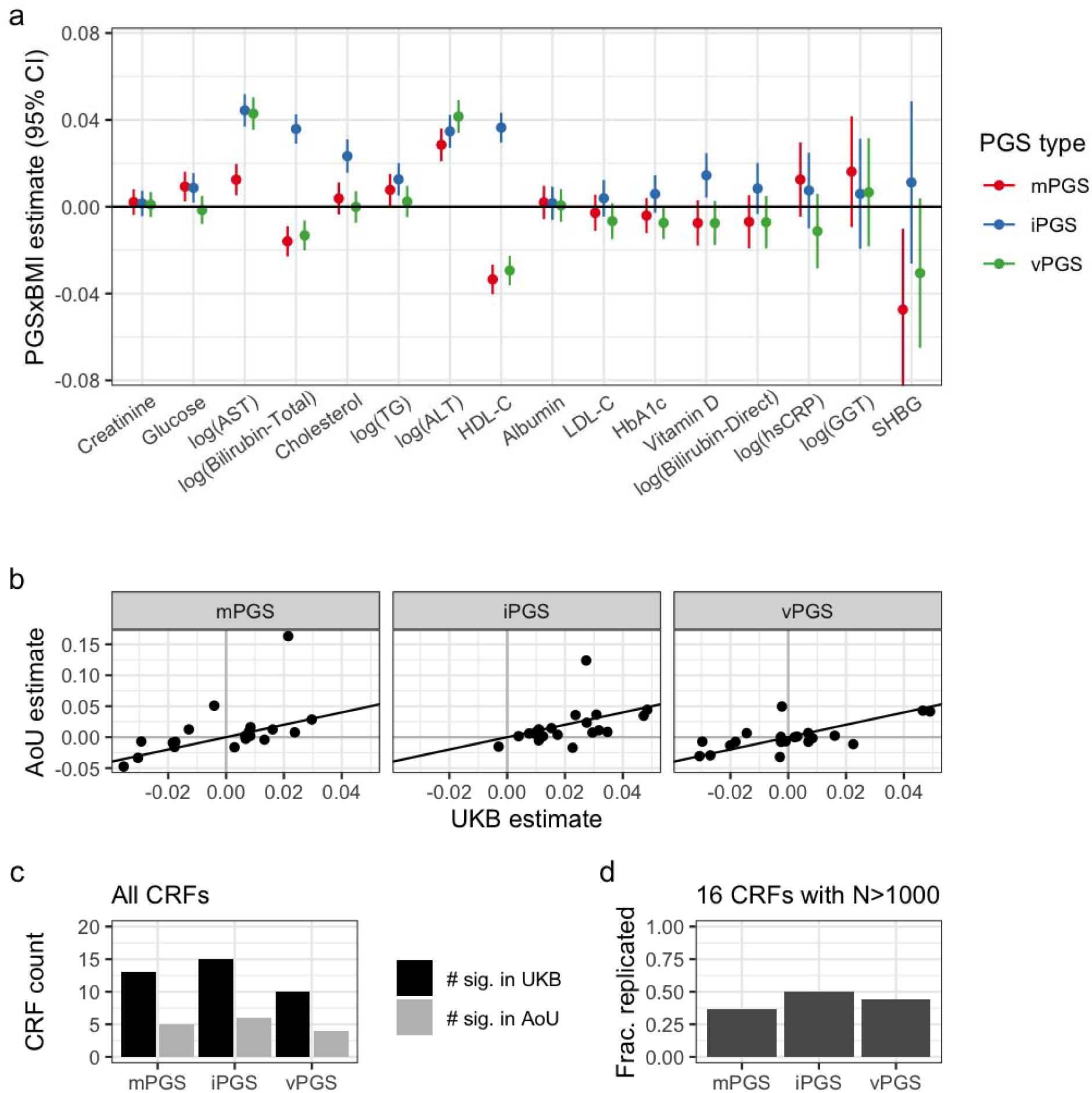
Replication results from All of Us. (a) Standardized interaction estimates (units of *SD*_*Y*_ / *SD*_*PGS*_ / *SD*_*BMI*_) are plotted for each combination of CRF (*x*-axis; ordered by decreasing sample size) and PGS type (colors). Results are only shown for CRFs with sample size greater than 1,000. (b) Standardized AoU interaction estimates plotted against UKB estimates for the same PGS type (panels) and CRFs (individual points). (c-d) Replication of significant UKB interactions (black) in AoU (gray). *Y*-axis indicates the number of CRFs with significant interactions, either from all available CRFs (c) or only those with N>1,000 samples available for replication in AoU (d). Counts for AoU in gray are for only those CRFs that were significant in UKB.

The ancestral and ethnic diversity of the AoU dataset provided an opportunity to not only explore PGS generalizability in a substantially different population from the European-focused UKB, but also test their performance in specific population strata. Though existing results suggest that P&T-based PGS generated in the European-enriched UKB do not generalize as well to non-European populations^27^, we did not see a major difference in PGS×BMI interactions in the pooled (37% non-European) versus European-only subsets (Supp. Fig. S8).

### Genetic modification of the BMI-ALT relationship

PGS performance was especially strong for two liver-related CRFs, alanine aminotransferase (ALT) and aspartate aminotransferase (AST), for which higher levels can indicate liver damage. For both, the iPGS and vPGS outperformed the mPGS in the UKB testing set (Fig. 3) and the AoU replication dataset (Fig. 4), with substantial consistency across multiple ancestry groups (Supp. Fig. S9). On this basis, we further interrogated the performance and biology of these PGS, focusing on ALT due to its greater specificity for liver function.

In both cohorts, we observed a strong positive correlation between BMI and log(ALT) (Fig. 5a). Likewise, iPGS-stratified analysis demonstrated the increasing magnitude of the BMI-log(ALT) association in step with the iPGS, especially in its highest deciles; this was consistent in both cohorts despite a weaker overall BMI-log(ALT) association in AoU (Fig. 5b). As another, more clinically applicable angle on this effect heterogeneity, covariate-adjusted BMI-log(ALT) associations were substantially stronger in the top iPGS decile compared to the remaining 90% of the population (Fig. 5c). The relative magnitude of this increase was much larger in AoU (72%) than UKB (27%), which can be traced to the smaller general BMI-log(ALT) effect size in AoU.

**Figure 5:**
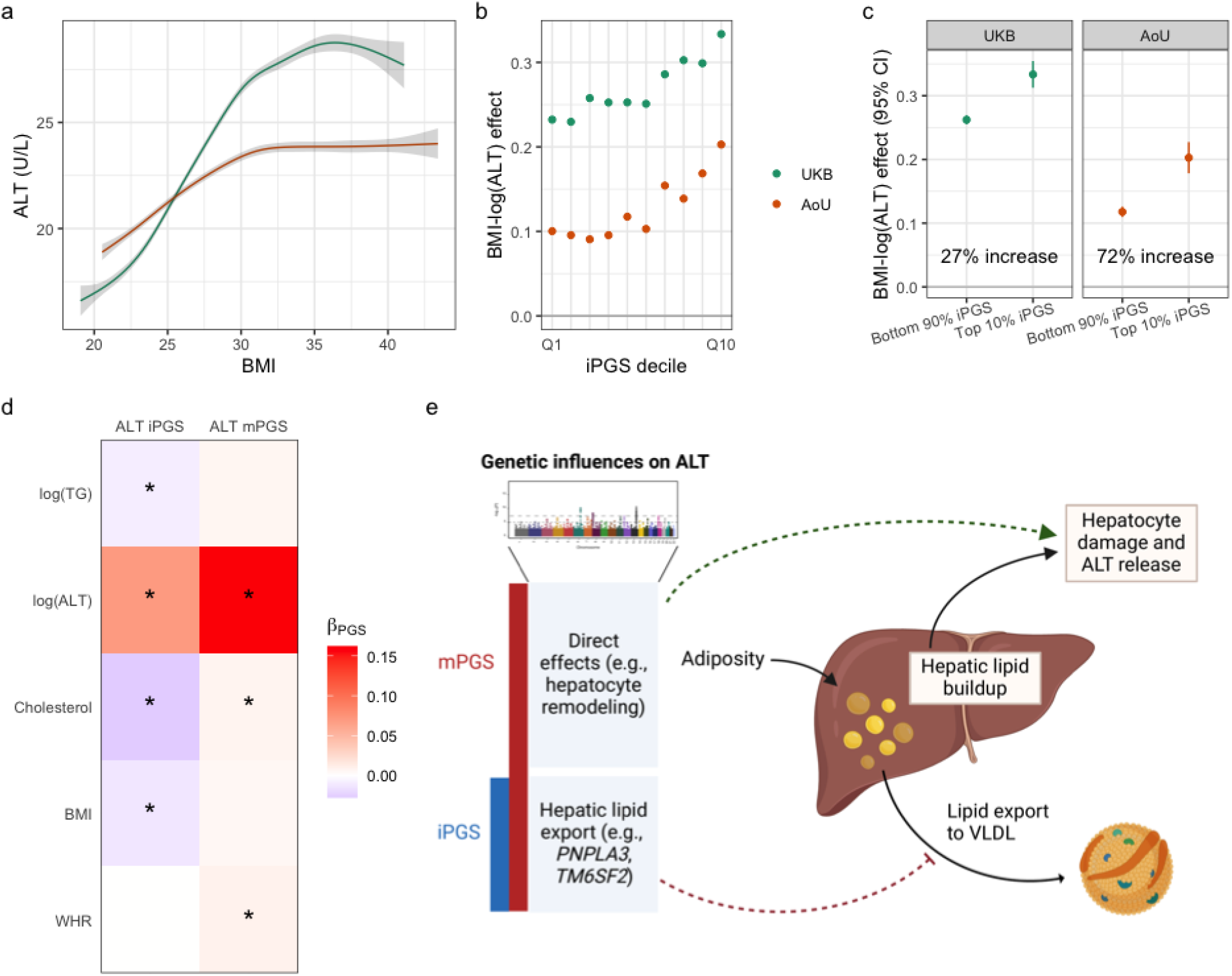
Exploration of the genetic modification of the BMI-ALT relationship. (a-b) Smooth spline curves (shrunken cubic spline) of the BMI-CRF relationship with 95% confidence intervals in gray. Curves are based on samples between the 5^th^ and 95^th^ percentiles of the BMI distribution. (b) Regression effect estimates of BMI on log(ALT) (*y*-axis), stratified by PGS decile (*x*-axis). (c) Regression effect estimates of BMI on log(ALT) (*y*-axis), stratified by a cutoff for “high” iPGS values (colors correspond to iPGS bin; threshold displayed on *x*-axis). (d) Heatmap shows regression coefficients linking the mPGS and iPGS for ALT to biologically relevant CRFs and adiposity measures. Stars correspond to associations with *p* < 0.05. (e) Diagram shows how the mPGS and iPGS capture different components of the genetic effect on ALT, with only a subset modifying the causal effect of BMI/adiposity.

Moving beyond their regression performance, we make several observations about the biological mechanisms captured by the iPGS for ALT compared to the mPGS. At the optimized P&T *p*-value threshold (*p* < 5×10^−8^), the iPGS contains signals from only a handful of genomic regions (11 variants in 8 loci; Supp. Table S9), all of which are within 100kb of one of the 311 variants composing the mPGS. These eight loci correspond to genes that are well known in liver-related diseases like metabolic disease-associated steatotic liver dysfunction (MASLD), such as *PNPLA3* and *TM6SF2*^28^. The iPGS and mPGS also associated differently with other CRFs. For example, the mPGS was positively associated with total cholesterol (TC) and triglycerides (TG; nonsignificant), suggesting that it captures effects related to broader metabolic dysfunction (associations shown in Fig. 5d). In contrast, the iPGS was negatively associated with TC and TG, consistent with a mechanism in which higher BMI leads to hepatic lipid buildup and genetic factors (encoded by the iPGS) reduce the liver’s ability export these lipids within lipoproteins (Fig. 5e). This phenomenon is known to involve both *PNPLA3* and *TM6SF2* and explains the discordant effects of some genetic variants on liver disease versus coronary artery disease^28,29^. Thus, without explicitly leveraging external variant or pathway annotations, the iPGS for ALT ultimately “selected” BMI-interacting genetic factors from the set of all biological pathways related to liver stress.

To support the above conclusions, we conducted gene set enrichment analysis using the full set of ALT GWAS and iGWAS summary statistics. Indeed, the Reactome gene sets most enriched for iGWAS signal were related to lipoprotein assembly and export (e.g., Plasma Lipoprotein Assembly, *p*_enrichment_ = 2.7×10^−6^; Supp. Table S10). In contrast, this pathway had minimal enrichment for GWAS signal (*p*_enrichment_ = 0.11), which instead was enriched for pathways relating to cellular remodeling and response to stress (e.g., Rho GTPase Cycle, *p*_enrichment_ = 2.6×10^−6^).

## Discussion

Here, we conducted a comprehensive exploration of PGS optimized for the detection of polygenic G×E, with implications for response to changes in physiology, clinical treatments, or lifestyle. Our simulations suggested that the iPGS approach is often most powerful, but that the mPGS or vPGS may be more effective in some cases, depending on the distribution and measurement accuracy of the exposure. Our applied analysis revealed an influence of genetics on the BMI-CRF relationship across a broad range of CRFs (most notably markers of liver stress), with iPGS most frequently capturing the strongest interactions.

The most important contribution from our simulations was manipulation of the exposure distribution and measurement accuracy. Most biological quantities are nonnegative, whether molecular (e.g., concentrations of some factor in blood) or behavioral (e.g., physical activity levels), and this should be accounted for when conducting G×E-focused simulation studies. As previously described, the exposure distribution in the underlying data-generating model is a critical factor in interpreting G×E interaction results and directly impacts the degree to which tests of marginal genetic effects (as used in the mPGS in this study) will capture G×E relationships^23^. We note that the exposure measurement quality, while a substantial concern in general, is less concerning for the BMI exposure used in this study’s applied analysis.

Based on theoretical expectations and our simulation results, we can make some preliminary statements comparing these PGS approaches for G×E detection. The mPGS, which leverages the highest-powered underlying statistical test (compared to G×E or vQTL single-variant tests) will be the most powerful when the true underlying exposure has a mean that is far from zero^23^. The mPGS performance will also track with the underlying correlation between genetic main and interaction effects, as demonstrated empirically in our UKB results (Fig. 3e,f). The iPGS approach will likely be optimal for detecting PGS×E in many cases, given the alignment between its underlying variant-specific statistical test (G×E interaction) and the ultimate PGS×E interaction test of interest. Importantly, its performance depends on having sufficient statistical power in these underlying single-variant interaction tests (which in turn depends on factors like sample size and measurement error). Finally, the vPGS will be most effective in particular cases involving very poor exposure measurement, which disproportionately hurts the performance of the G×E test/iPGS approach that directly uses measured exposure values. Though not included in these simulations, we note that vPGS performance will also degrade with the number of overall genetic effect modifiers (genetic and non-genetic), since vQTL tests are not specific to the exposure in question.

Our applied analysis considered genome-wide genetic modification of the BMI-CRF relationship. This builds on previous observations of polygenic interactions with adiposity: stratification by BMI has been shown to improve the performance of PGS for type 2 diabetes (allowing increased contribution of beta cell-related pathways in low-BMI individuals ^20^) and the inclusion of interactions with waist-hip ratio (a related adiposity measure) modestly improves the predictive performance of PGS for blood biomarkers^11^. Here, we explored a broad set of CRFs with a specific focus on comparing the performance of the three PGS types. In this study, the iPGS approach showed the most consistent performance across CRFs, matching expectations as described above. The less consistent performance of the vPGS may reflect the fact that BMI is measured well, meaning that there is less relative benefit compared to the iPGS. However, in many cases, the vPGS detected equally strong interactions to the iPGS (e.g., for ALT and AST) and mPGS (e.g., for SHBG and direct bilirubin). Its interaction effects tracked most closely with the mPGS, which is unsurprising given the close relationship between genetic mean and variance effects^15^. We also note that vPGS may have applications beyond capturing G×E, such as predicting within-individual variability over time^16^.

Our mPGS results are consistent with a series of recent studies supporting amplification as the primary mode of polygenic G×E^4,30,31^. In this framework, disease-associated genetic predisposition and disease-associated exposures are synergistic in increasing risk. We note that these studies primarily leverage standard PGS, comparable to our mPGS analyses. Indeed, we see positive mPGS×BMI interactions for CRFs with positive disease and BMI associations and negative interactions for CRFs with negative associations (i.e., the mPGS and BMI are synergistic; Supp. Fig. S4). Importantly, this does not mean that the amplification model is correct for every contributing genetic locus.

As noted above, the choice to include an mPGS main effect in iPGS (or vPGS) interaction models is not expected to impact type I error. Rather, it may reduce the standard error of PGS×E estimates by explaining additional variability in the outcome, though our primary UKB results using the iPGS did not meaningfully change when including this adjustment (as shown in Supp. Fig. S6). In general, this choice of adjustment strategy may differ depending on the goal of the analysis. For many studies including PGS×E interactions, the goal is enhancing outcome prediction beyond solely standard PGS (as is most commonly explored in the current iPGS literature). In that case, it is naturally important to include the main effect PGS (which will typically explain much more variance than the interaction effect, based on existing results from whole-genome variance components approaches^32^). However, in cases like the present study, for which the evaluation of a PGS×E effect is of primary interest, adjustment for a separate PGS (beyond that being evaluated for interaction) may complicate interpretation if it is correlated with the focal interaction term of interest.

We saw particularly strong interactions of BMI with the ALT and AST iPGS, reinforcing known variant-specific findings from a European ancestry subset of the UKB^33^. We conducted a more in-depth investigation of the ALT iPGS to explore potential biological mechanisms. Compared to the associated mPGS, iPGS had specific contributions from alleles hindering the export of hepatic lipids (known to be increased in obesity^24,34^) in the form of lipoproteins. This mechanism was supported by inverse associations of the iPGS with circulating TG and TC as well as by pathway enrichment for lipoprotein assembly and export. In contrast, associations between the ALT mPGS and these traits were positive; this is a more intuitive relationship given that general metabolic dysfunction increases both liver stress and circulating lipid levels. As referenced above, this contrast reveals a key insight about the iPGS strategy: from the overall genetic architecture of a trait, it has the capacity to naturally “select” subsets of variants related to specific biological mechanisms interacting with the exposure of interest. Our results further converge with findings derived from more hypothesis-driven “partitioned PGS” constructed based on the consistency of variant effect directions with liver fat versus circulating TG^35^.

This study represents the first effort to directly compare the performance of genetic main, interaction, and variance PGS within a unified analysis pipeline for detecting interactions. It is strengthened by the incorporation of best practices from existing PGS pipelines and genome-wide studies for each of the relevant approaches. However, there remains uncertainty as to the optimal way to generate and test these response-focused PGS. Especially for vQTL studies, it is not clear which statistical framework is optimal for single-variant studies nor associated vPGS (though recent studies are beginning to make the necessary comparisons^36^). An additional limitation is that our applied investigation uses only BMI as an exposure and cardiometabolic biomarkers as outcomes. Different exposures, such as lifestyle-related variables or pharmaceutical drugs might have substantially different “response genetic architectures”, and even within the domain of adiposity, BMI is an imperfect measure that doesn’t capture body fat distribution and reflects other factors such as muscle mass^37^. We intentionally selected continuously-valued CRFs for this study to accommodate the inclusion of vQTL tests, but the iPGS strategy can be straightforwardly applied to binary outcomes as well with some additional methodological considerations^22^. Finally, the three score types tested here are not exhaustive; for example, machine learning-based approaches can model the sensitivity of exposure-disease relationships to genetic factors^38^.

In summary, our study contributes to the accumulating literature establishing polygenic contributions to G×E. Our results support and expand existing observations about iPGS performance^5,22^ and the amplification model^4,30^ and show that genetic factors meaningfully alter the relationship between adiposity and cardiometabolic risk. These findings indicate the potential of genetic scores to contribute to more personalized chronic disease prevention strategies.

## Methods

### Simulation study

A simulation study was conducted to validate existing results in the literature, explore additional conditions under which these scores might have inflated type I error, and compare their statistical power for the detection of PGS×E interaction under varying assumptions about the exposure distribution and measurement characteristics. The simulation workflow is depicted in detail in Supp. Fig. S1. We summarize it here:

First, a single genotype dataset was generated for *N* = 10,000 samples and M = 100 independent variants, with genotype values drawn from a binomial distribution and minor allele frequencies (MAFs) drawn from a uniform distribution between 0.05 and 0.5.

Next, for each simulation scenario (defined by a specific set of parameter specifications), a set of P phenotypes were generated. To accomplish this, P random, standard normally-distributed exposures E were first simulated, with possible contribution from the simulated G based on a specified variance explained 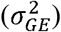. A corresponding series of P standard normal phenotypes were then generated with contributions from G and E main effects, a G×E product term, and additional random error to produce a final outcome variable with variance one. Parameters specified included: variance explained by exposure main effects 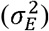, genetic main effects 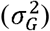, and their interaction 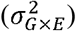,

After phenotype simulation, a random 70% of each simulated sample was assigned to the training set, with the remaining 30% assigned to the testing set. For each of these P phenotypes, a “genome-wide” set of results (for each statistical test) was generated in the training subset by performing a series of statistical tests for marginal effects (estimating *β*_*G*_), interaction effects (estimating *β*_*GXE*_), and variance effects (estimating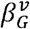). Marginal and interaction effects were generated by their respective standard linear models in R, while variance effects were estimated using the DRM method, which regresses absolute deviations from genotype-specific median values on additively-coded genotype values^15^. These summary statistics were converted into PGS weights via simple thresholding: regression estimates were used directly for variants with *p* < 0.05, and weights were otherwise set to zero. Each of the three PGS were then calculated as weighted sums based on these weights. Finally, these PGS (three per phenotype vector) were tested for PGS×E interaction using a significance threshold of *p* < 0.05 to determine type I error (for 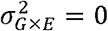) and power (for 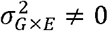).

A few extensions of this base simulation framework were used to expand the exposure component. A gamma-distributed E (shape and rate parameters equal to one) was used to explore the effect of a nonnegative E. In this case, when G-E correlation was specified based on a nonzero 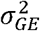 parameter, the betas for each G were generated as usual, but then exponentiated and multiplied by the gamma-distributed E, before scaling the E back to variance one. Power simulations with a normally-distributed E also tested the effect of measurement error, for which a parameter (*ICC*_*E*_) specified the amount of variance in the measured E explained by the “true” E, i.e., the amount of additional noise introduced.

Simulations and all subsequent analyses were conducted using R versions 4.1 and 4.2^39^ except where otherwise noted.

### UK Biobank cohort

This work was conducted under a Not Human Subjects Research determination for UKB data analysis (NHSR-4298 at the Broad Institute of MIT and Harvard), under UKB application 277892. UKB is a large prospective cohort with both deep phenotyping and molecular data, including genome-wide genotyping, on over 500,000 individuals ages 40-69 living throughout the UK between 2006-2010^40^.

Genotyping, imputation, and initial quality control on the genetic dataset have been described previously^41^. Work was conducted on genetic data release version 3, with imputation to both Haplotype Reference Consortium and 1000 Genomes Project (1KGP). For sensitivity analyses in ancestry-specific data subsets, genetic ancestry labels were retrieved from the Pan-UKBB project^41^.

Body mass index (BMI; kg/m^2^), the primary exposure of interest, was collected from assessment center anthropometric measurements. As outcomes, we focused on 20 serum biomarkers related to cardiovascular disease and metabolism, including but not limited to lipids, liver enzymes, glycemic parameters, and kidney function markers (see Supp. Table S2). Blood samples were collected at the baseline visit for the majority of participants, and specific biomarkers were measured using colorimetric, enzymatic, and immunoassays (details available at: https://biobank.ctsu.ox.ac.uk/crystal/crystal/docs/serum_biochemistry.pdf).

We excluded individuals that had withdrawn consent by the time of analysis excluded as well as those with diabetes, coronary heart disease, cirrhosis, end-stage renal disease, cancer diagnosis within one year prior to their assessment center visit, or who were pregnant within one year of the assessment center visit. Cholesterol, LDL-C, and Apolipoprotein B were also adjusted for statin use using methods described previously^42^: in individuals with self-reported use of a statin medication, each of these biomarkers was divided by an adjustment factor (0.749, 0.684, and 0.719, respectively) that had been empirically estimated by Sinnott-Armstrong and colleagues^21^ in the same population. After these adjustments, a subset of highly skewed biomarkers was log-transformed (see Supp. Table S2) and outliers (greater than 5 standard deviations from the mean) were set to missing for BMI and all biomarkers. Finally, we further subset to a group of unrelated samples used for genetic principal components (gPCs) analysis during central genetic data preprocessing^43^.

After all phenotype preprocessing steps, the unrelated, multi-ancestry UKB sample was randomly subdivided into three groups: training (70%), optimization (10%), and testing (20%). This split devotes a majority of the sample to the generation of genome-wide summary statistics, which contribute to PGS performance and out-of-dataset generalizability, and follows similar splits used in PGS analyses^21,44^. Due to the substantial correlation between the 20 CRFs, we calculated a smaller number of “effective” biomarkers in the training set using a PCA-based approach we have previously deployed for blood biomarkers in this dataset^26^.

### Genome-wide models

For each biomarker of interest, three statistical models were run genome-wide in the UKB training set to generate summary statistics that would inform subsequent PGS development. Each was run on common variants (MAF >1%) with imputation INFO score greater than 0.5.

#### i. Main effects: genome-wide association study (GWAS)

The GWAS model is as follows:

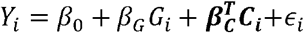

Where *Y*_*i*_ is the outcome for individual *i, G*_*i*_ is the genotype vector, *C*_*i*_ is a vector of covariates, and *ϵ*_*i*_ captures residual error. Covariates included sex, age, age, an age-by-sex product term, and 10 gPCs. This model produces *β*_*g*_ (genetic main effect) estimates and *p*-values for each variant. The GWAS models were run using the GEM program ^45^ with no exposure specified and model-based standard errors.

#### ii. Interaction effects: genome-wide interaction study (iGWAS)

The iGWAS model is a straightforward extension of the GWAS model:

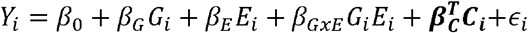

Where additional terms have been added for *E*_*i*_, the exposure, and its product term with *G*_*i*_. Covariates matched those from the GWAS, with the addition of exposure-by-gPC product terms for each of the 10 gPCs (as found to be critical in pooled ancestry interaction analyses^46^. The key estimate of interest from this model is *β*_*GxE*_ (the interaction effect), rather than *β*_*G*_. The iGWAS models were run using GEM with mean-centered BMI as the exposure and robust standard errors.

#### iii. Variance effects: genome-wide variance study (vGWAS)

Variance-quantitative trait locus (vQTL) analysis quantifies genetic effects on trait variability (rather than mean). This analysis doesn’t directly model interactions, but may nonetheless have greater power in some cases to detect variants supporting interaction with environmental exposures. For example, the deviation regression model (DRM) from Marderstein and colleagues^15^ is:

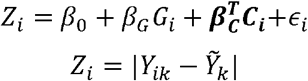

Where *k* indexes the genotype group corresponding to individual *i* and 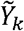 is the median phenotype value in genotype group *k*, such that *Z*_*i*_ represents the individual’s absolute deviation from the genotype-specific median. For computational efficiency and statistical robustness, the statistical model used in the applied UKB analysis is the quantile integral linear model (QUAIL)^16^. A standard quantile regression model follows:

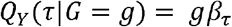

Where τ is the quantile and *β* _τ_ is the regression coefficient associated with that quantile. In this setup, *β*_1-τ_ −*β*_τ_ corresponds to the vQTL effect (i.e., the genotype effect on the quantile differs across lower versus higher quantiles). Aggregating information across quantiles results in the quantile-integrated model tested in the QUAIL program:

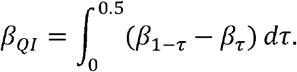

Which can be tested for genetic variants genome-wide (see Miao 2022 for more details), with these *β*_QI_ (denoted moving forward as 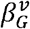, indicating “variance”, for clarity), being the primary estimates of interest)^16^.

### Polygenic score generation and optimization

A series of mPGS were generated for each biomarker of interest using a basic P&T strategy as implemented in the PRSice-2 program^47^. Inputs included GWAS summary statistics (effect estimates and *p*-values) and an LD reference panel consisting of a random 20,000 individuals from the UKB. For pruning within PRSice-2, a grid of *p*-value thresholds of 0.05, 0.01, 0.005, …, 1×10^−7^, 5×10^−8^ was used, along with a clumping radius of 250kb and r^2^ threshold of 0.1. Ambiguous variants (A/T or C/G) and those with duplicated rsIDs in the original UKB annotation file were excluded during the PGS development step, prior to pruning.

Though the standard PRSice pipeline includes PGS *p*-value threshold optimization using linear regression models in the target dataset, we did not use this functionality here since the goal was to optimize for the detection of interaction rather than main effects. Instead, this optimization was performed separately, using a held-out optimization subset of the UKB. Specifically, mPGS corresponding to each value of the threshold were included in separate regression models including main and exposure interaction effects:

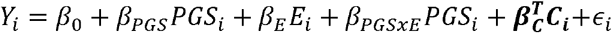

Covariates for these optimization regressions were identical to those from the iGWAS (i.e., including 10 BMI-by-gPC product terms). The optimal threshold chosen to minimize the *p*-value of the estimated interaction effect, *β*_*PGSxE*_. Finally, the PGS corresponding to the optimal *p*-value threshold was evaluated using the same regression model in the fully held-out testing subset.

PGS generation, optimization, and evaluation proceeded similarly for the iPGS (using *β*_*GxE*_ estimates) and vPGS (using *β*_*v*_ estimates).

### Additional follow-up analyses

Genetic correlations between genetic main and interaction estimates from iGWAS were estimated using bivariate LD-score regression (LDSC)^48,49^. For each CRF, genetic main effect and interaction effect estimates and *p*-values (using robust standard errors) were retrieved from the same set of iGWAS summary statistics. LDSC was then run using a European-ancestry linkage disequilibrium reference dataset from the 1000 Genomes Project (https://alkesgroup.broadinstitute.org/LDSCORE/). Given the sensitivity of genetic main effect estimates to the centering of the interaction exposure^23^, we reiterate that BMI was mean-centered prior to the iGWAS feeding these LDSC runs.

Variants included in selected PGS were annotated to genes using ANNOVAR^50^ (version 2018-04-16; based on genome build GRCh38).

ALT-specific GWAS and iGWAS results were subject to enrichment analysis to prioritize gene sets with enrichment of signal in the surrounding genetic region. *P*-values from the associated genome-wide summary statistics were used as input to the MAGMA program^51^, using the same LD reference panel as above and gene regions defined as 2kb upstream to 1kb downstream of the gene limits based on the NCBI database (GRCh37). Gene sets from the Reactome pathway collection^52^ were downloaded from mSigDB^53^.

### All of Us cohort

The AoU cohort contains data from over 413,000 participants, of which more than 245,000 have genetic data available from whole-genome sequencing. AoU operates under a “data passport” model in which project-specific IRB approval is not needed for the analysis of de-identified data. AoU research, as Participants were assigned to specific genetically-inferred ancestry groups, including African/African American (AFR), American Admixed/Latino (AMR), East Asian (EAS); European (EUR) and South Asian (SAS). Genetic principal components (gPCs) were available from central genotype preprocessing. Basic covariates, including sex at birth and age [determined from date of birth], were derived from survey responses.

BMI (concept ID: 3038553) was available as measured from outpatient settings (visit occurrence concepts: “Outpatient Visit” or “Office Visit”). To account for multiple measurements, all non-missing BMI values were averaged within each person. Outliers (greater than 5 interquartile ranges [IQRs] from the median) followed by values less than 10 were removed.

Blood biomarkers were chosen to match the 20 analyzed in the UKB (associated concept names provided in Supp. Table S6). They were retrieved as measured in outpatient settings (visit occurrence concepts: “Outpatient Visit”, “Office Visit”, or “Laboratory Visit”) and filtered for measurement using relevant units (Supp. Table S6). All valid biomarker values were averaged within each person. Finally, negative values were removed, zero values were imputed with half of the minimum non-zero value, and outliers (greater than 5 IQRs from the median) were removed.

PGS were calculated in AoU based on weights determined and optimized in the prior UKB analysis. Genotypes for relevant variants were retrieved from whole-genome sequencing (ACAF threshold callset; v7.1) using the Hail program^54^, based on chromosomal location, splitting multi-allelic variants using the split_multi_hts() function. PGS weights were harmonized by (1) flipping the sign of the PGS weight when the counted and non-counted alleles were the reverse of that from the UKB, and (2) dropping variants that were unavailable or for which alleles did not match. Score calculation was run using the “-- score” function from PLINK2^55^. For computational tractability, for the few scenarios in which the optimal score from UKB resulted in a very large number of variants included in the PGS, only the top 5,000 variants by *p*-value were used for PGS calculation.

AoU regressions mirrored those conducted in the UKB testing set, testing for PGS× BMI interaction as the primary estimate of interest while adjusting for sex at birth, age, age squared, 10 gPCs, and 10 gPC-by-BMI interaction product terms. Primary replication tests were performed in the full, multi-ancestry dataset, with PGS pre-adjusted for ancestry probabilities as previously described^56,57^. Ancestry-stratified sensitivity analyses were performed based on genetically inferred ancestry groupings^58^.

## Data Availability

No new genetic or phenotypic data have been generated for this study. The UK Biobank data, including genetic and phenotypic data, are under controlled access but can be obtained through application at https://www.ukbiobank.ac.uk/. UK Biobank will consider data applications from bona fide researchers for health-related research that is in the public interest. All of Us controlled tier data are available to authorized users on the Researcher Workbench (https://workbench.researchallofus.org/).

## Code Availability

Code supporting the conclusion of this manuscript can be found at: https://github.com/kwesterman/ipgs.

## Acknowledgements

We thank Andrew R. Marderstein for helpful feedback on the manuscript.

We gratefully acknowledge *All of Us* participants for their contributions, without whom this research would not have been possible. We also thank the National Institutes of Health’s *All of Us* Research Program for making available the participant data examined in this study.

Selected diagrams presented in this paper were generated using https://BioRender.com.

