## Supplementary figures for "Polygenic scores capture genetic modification of the adiposity-cardiometabolic risk factor relationship"

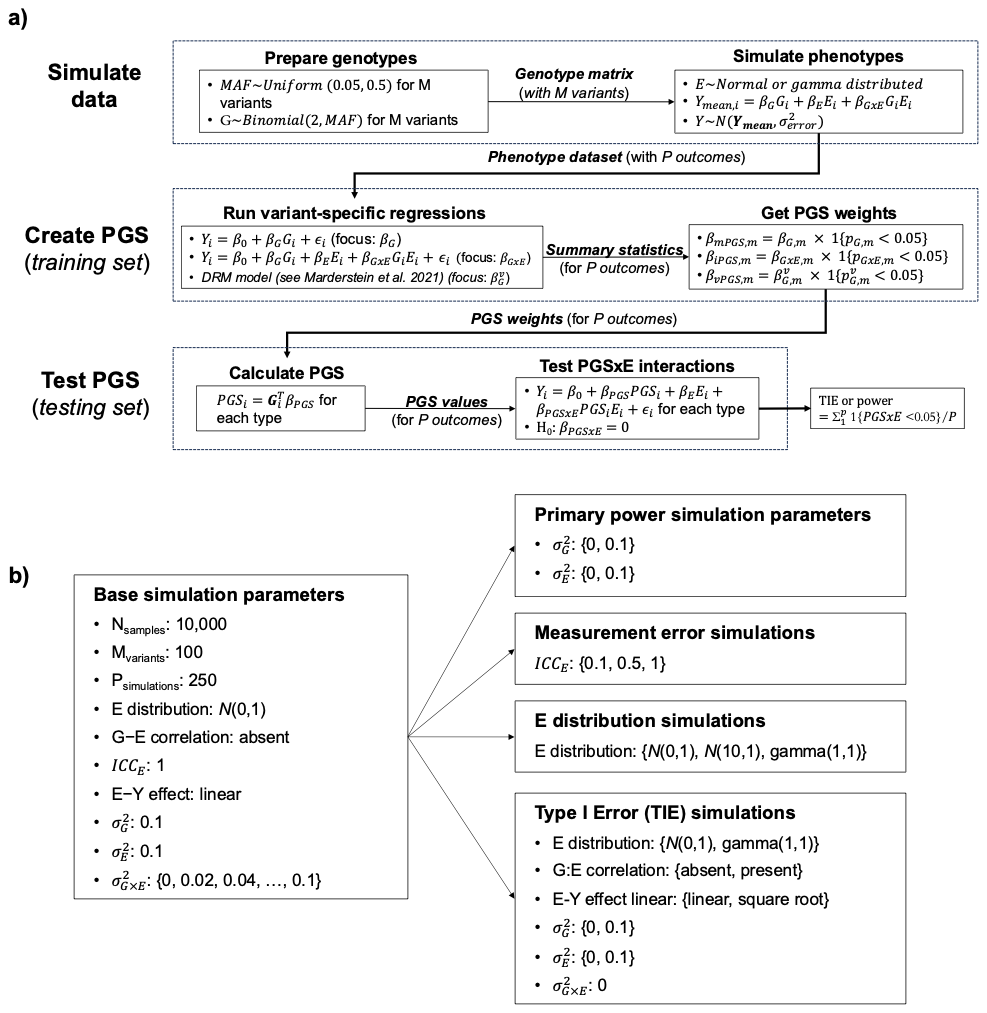
**Supplementary Figure S1:** Simulation study overview. (a) Simulation pipeline (see Methods for additional details). (b) Simulation parameters defining scenarios for type I error and power calculation. Base simulation parameters (left box) apply unless overridden by scenario-specific parameters (right boxes). G: NxM genotype matrix; E: NxP exposure matrix; Y: NxP outcome matrix.

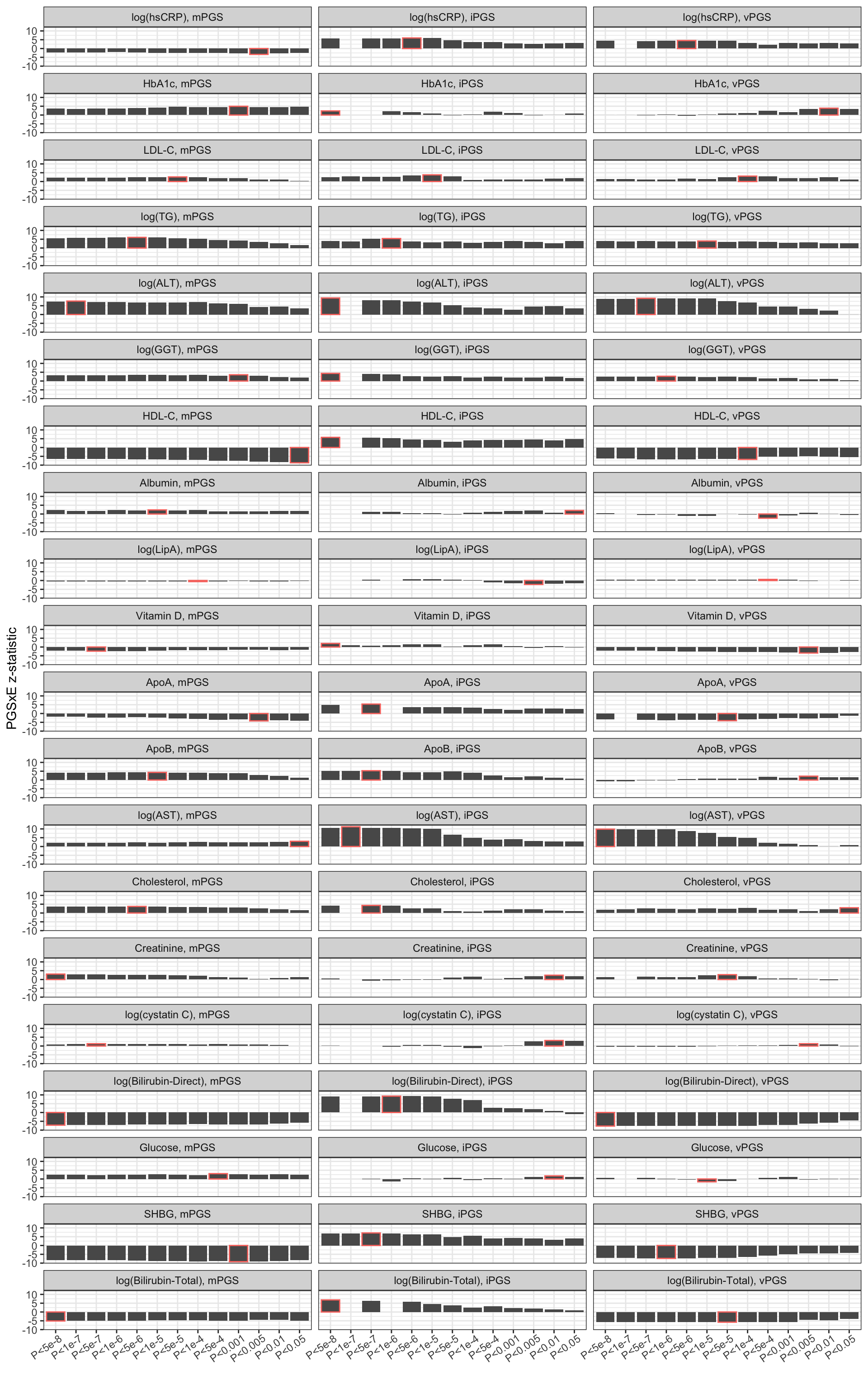
**Supplementary Figure S2:** Optimization of PGS for PGSxE interaction across all CRFs. Red boxes indicate the optimal *p*-value threshold selected (to maximize the magnitude of the xPGS×BMI z-statistic).

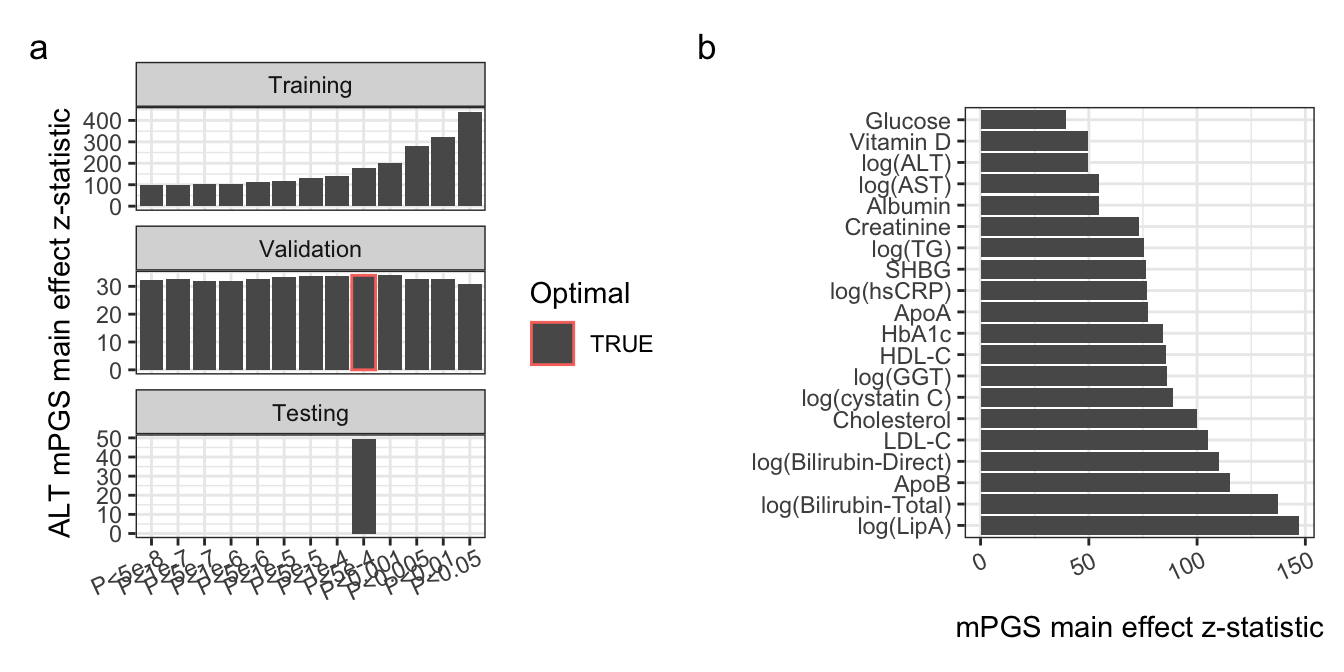
**Supplementary Figure S3:** Marginal effects of standard PGS as a positive control for the PGS development pipeline. (a) mPGS main effects in the training, optimization, and testing subsets. Red outline indicates the best-performing *p*-value threshold in the optimization subset. (b) Marginal mPGS effect z-statistics for each biomarker in the testing subset.

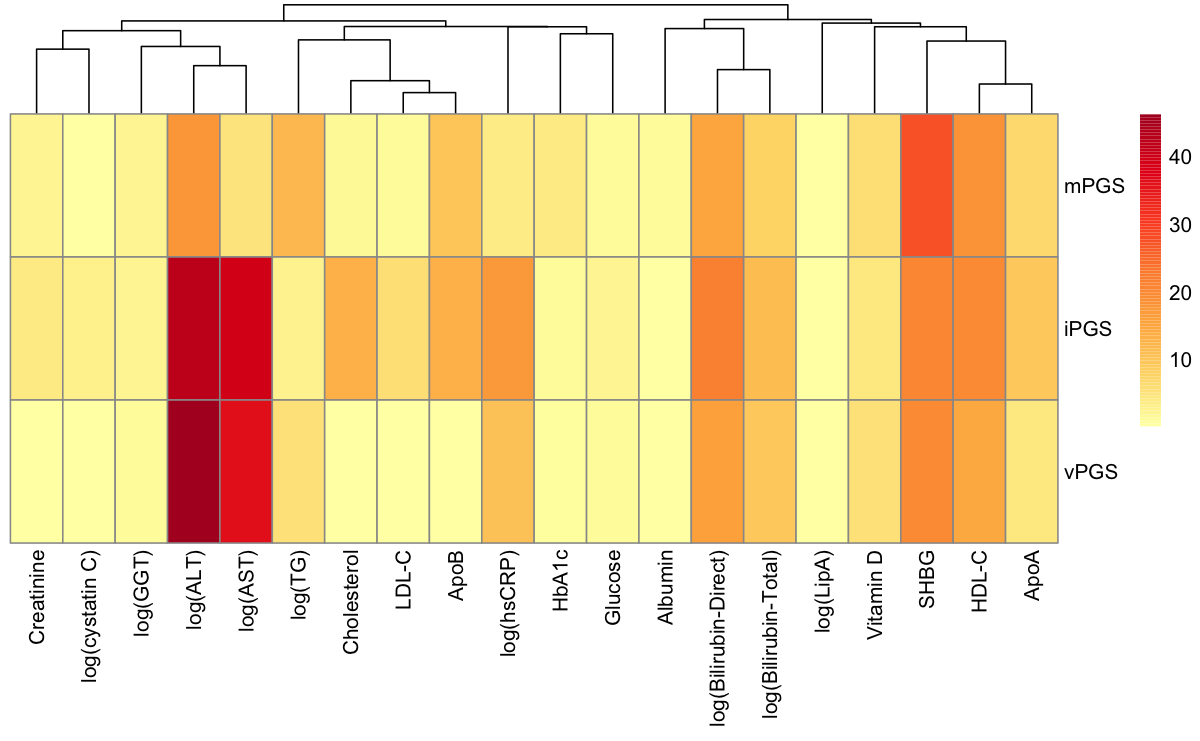
**Supplementary Figure S4a:** Heatmap of PGS performance (as measured by -log_10_(*p*_PGS×BMI_)) by CRF. CRFs are ordered based on a hierarchical clustering (Euclidean distance and complete linkage) of their values in the UKB training set after mean imputation of missing values.

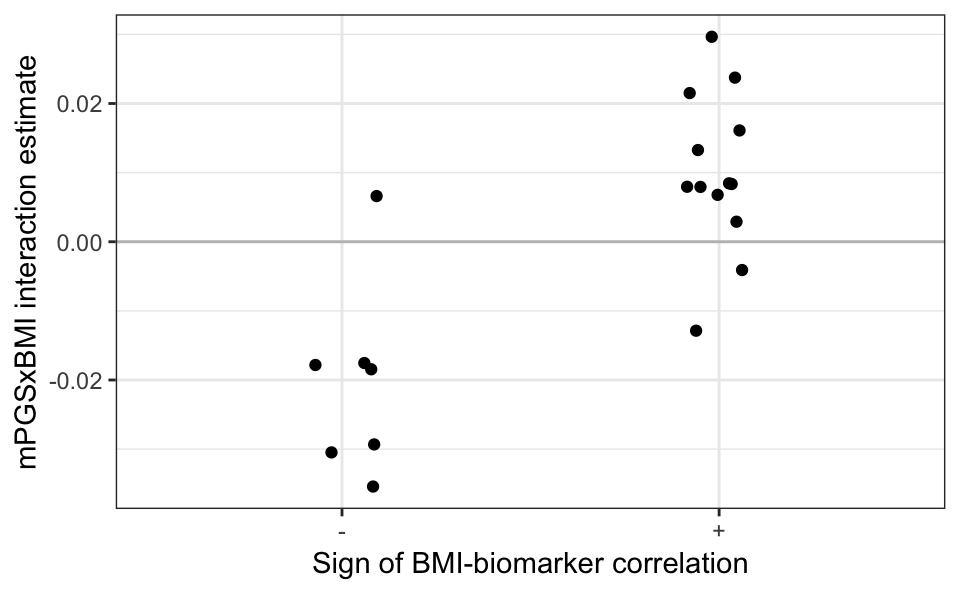
**Supplementary Figure S4b:** Relationship between negative mPGSxBMI estimates and directionality of the BMI-CRF relationship (with implications for the amplification model of GxE interactions). Interaction effects between the mPGS and BMI (*y*-axis) are plotted against the raw sign of the BMI-CRF correlation (*x*-axis).

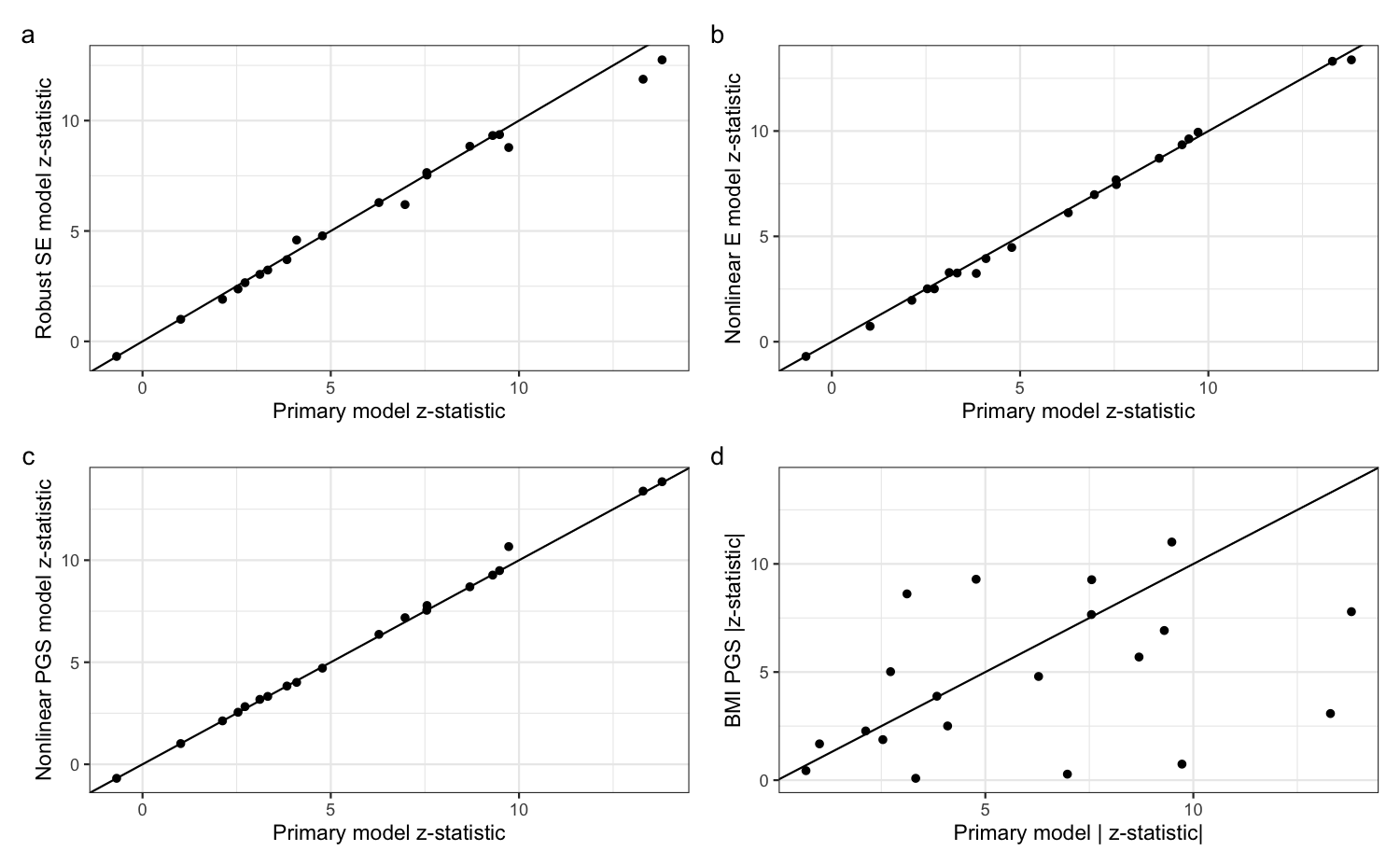
 **Supplementary Figure S5:** Sensitivity analyses addressing possible artifactual PGSxBMI in the UKB testing set. Plotted against primary iPGS model z-statistics for each of the 20 CRFs are z-statistics from models (a) using robust standard errors, (b) including a squared term for the BMI main effect, (c) including a squared term for the PGS main effect, and (d) replacing the iPGS with an mPGS for BMI (i.e., developed using BMI as the outcome, rather than the exposure), plotting absolute values of the z-statistics for . Solid lines denote *x* = *y*.

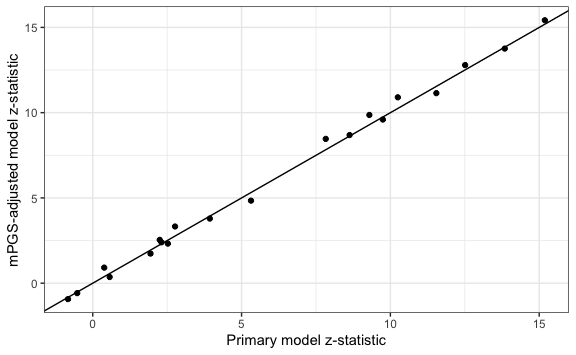

**Supplementary Figure S6:** Sensitivity analysis addressing the relevance of adjusting for mPGS to reduce the standard errors of estimates. For each of the 20 CRFs, z-statistics from iPGS interaction models are plotted, either from the primary model (*x*-axis) or additionally adjusting for an mPGS for that biomarker (*y*-axis). Solid line denotes *x* = *y*.

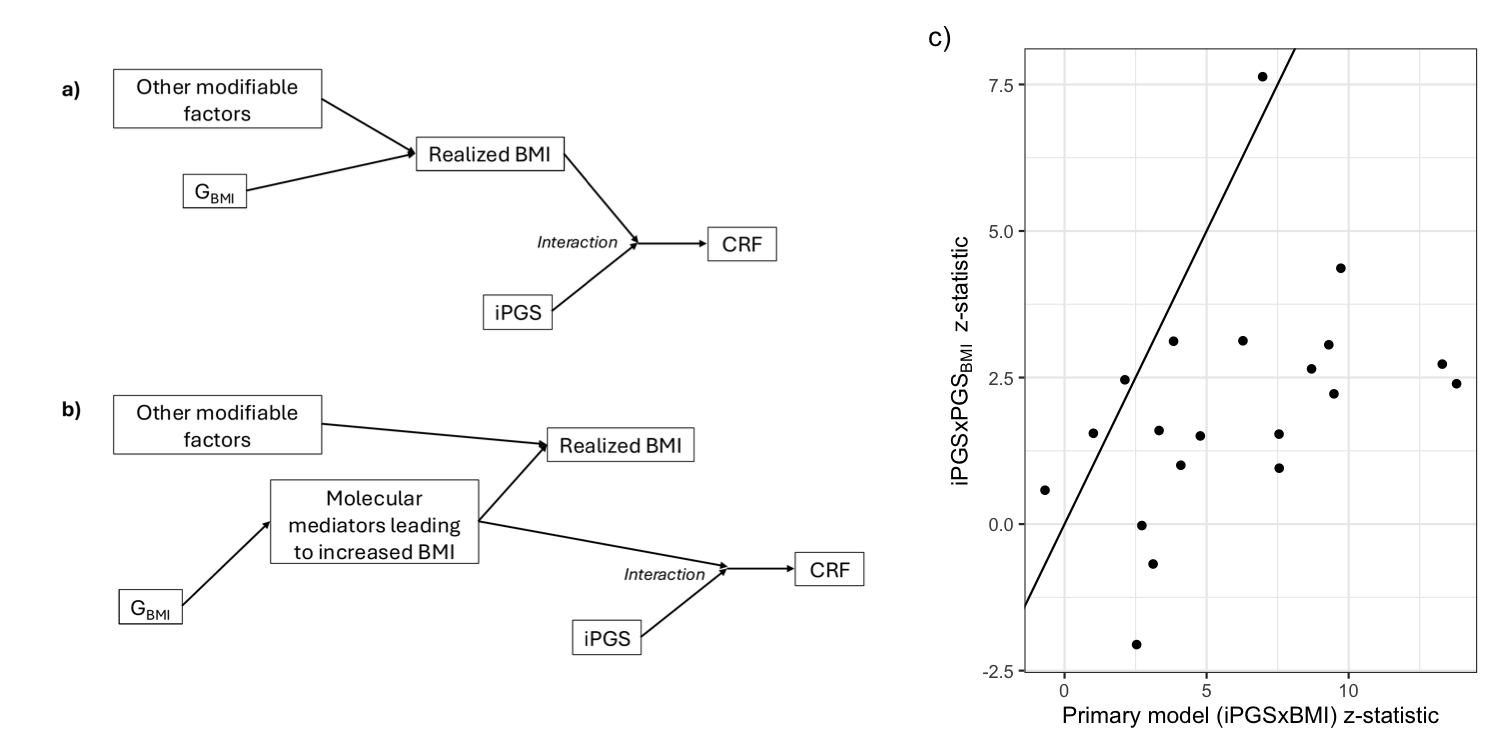
**Supplementary Figure S7:** Sensitivity analysis addressing the genetic underpinnings of BMI. a,b) Conceptual model describing the scenario in which replacing measured (“realized”) BMI with a PGS reflecting its genetic influences (here, G_BMI_) would or would not increase power for discovery of interactions with the iPGS. In (a), the iPGS interaction involves involves BMI itself, such that the upstream “cause” of BMI does not change the interaction strength. In (b), the iPGS interaction involves molecular mediators of the G-BMI relationship, such that replacing BMI with a genetic anchor might reveal a stronger interaction. c) Comparison of z-statistics across all CRFs from the primary iPGS tests (*x*-axis) or identical tests replacing BMI with an mPGS for BMI (*y*-axis). Solid line denotes *x* = *y*.

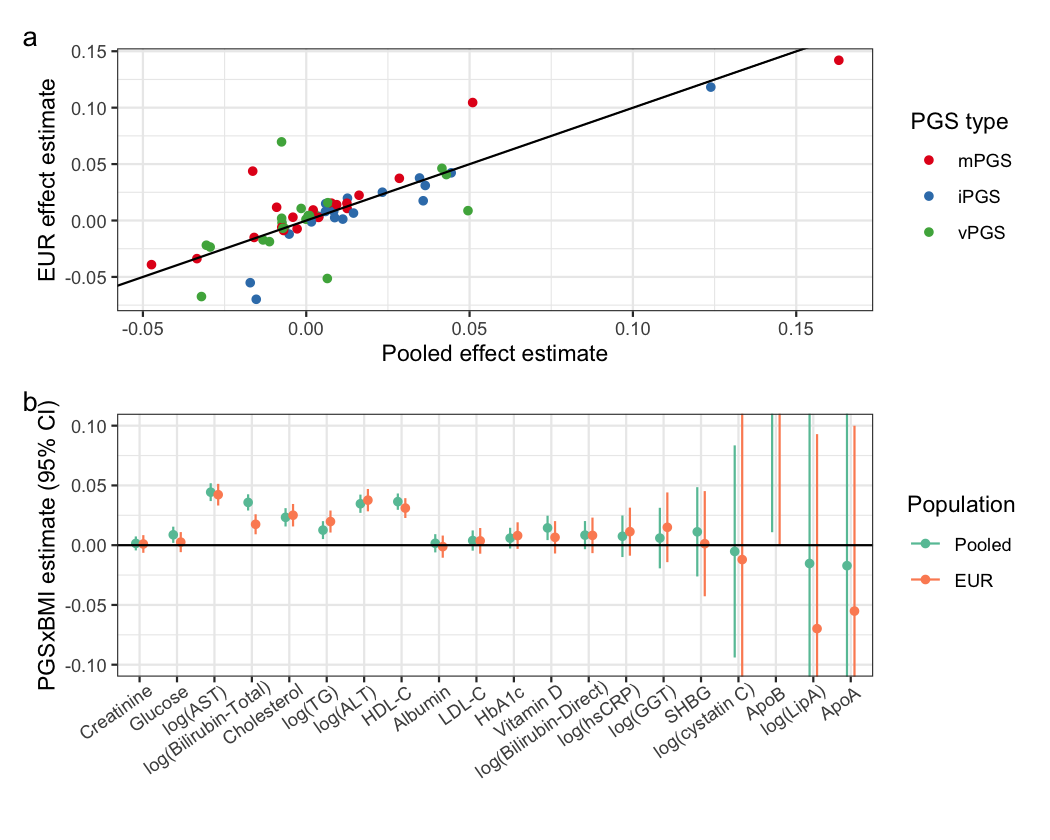
 **Supplementary Figure S8:** Consistency of PGSxBMI effects between the pooled and European-ancestry subsets of the All of Us dataset. (a) European ancestry-specific interaction effects are plotted against those from regressions using the full, pooled dataset (with gPC-based ancestry pre-adjustment). Colors correspond to PGS type. (b) Interactions effects specifically for the iPGS (*y*-axis) are plotted for each biomarker (*x*-axis), with colors corresponding to the population subset (pooled or European ancestry-only).

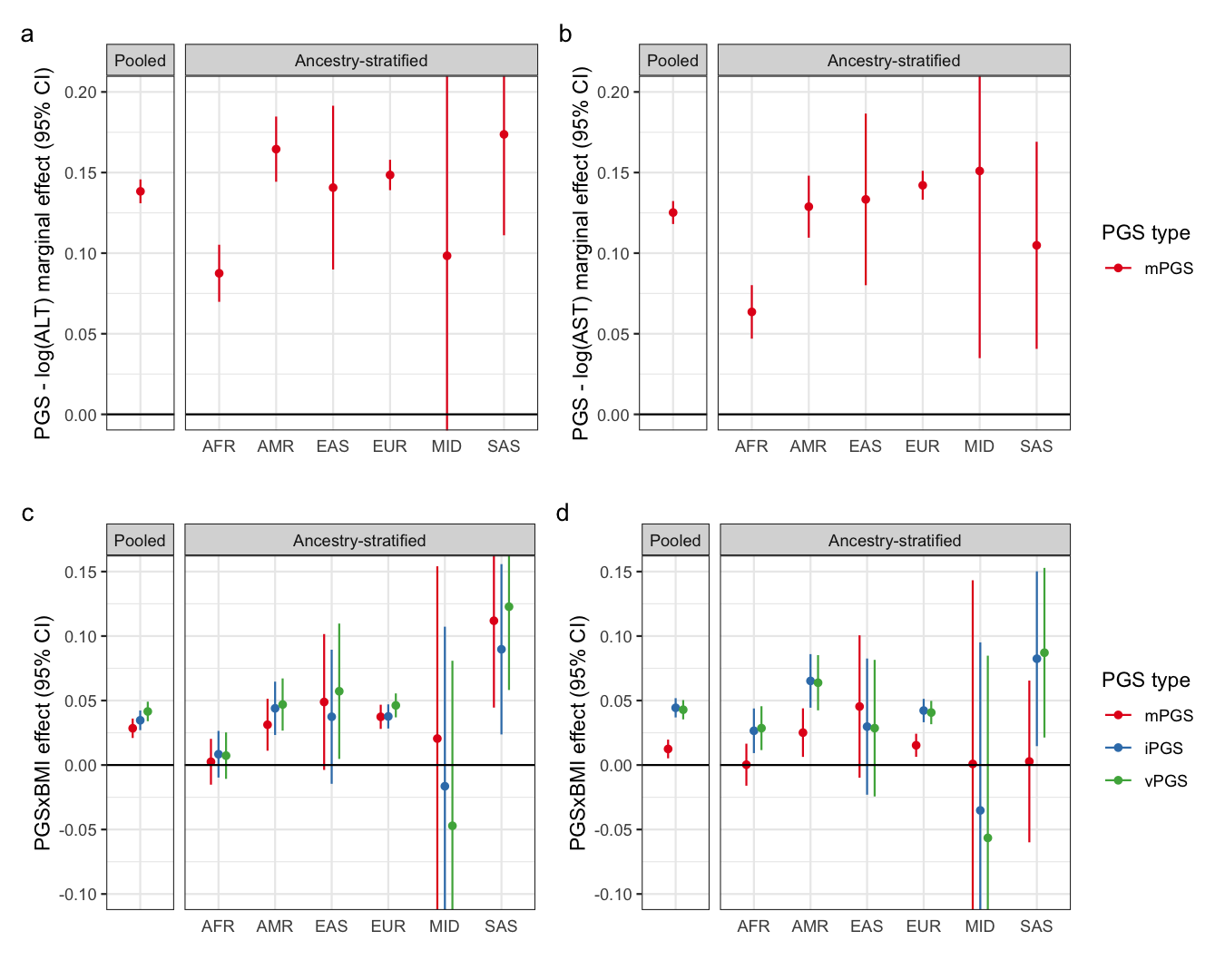
 **Supplementary Figure S9:** Ancestry-specific replication of marginal and interaction effects in All of Us. (a-b) Standardized marginal mPGS effects on log(ALT) (a) and log(AST) (b) in the full pooled dataset and genetic ancestry groups. (c-d) Standardized iPGS×BMI interaction effects on log(ALT) (c) and log(AST) (d) in the full pooled dataset and genetic ancestry groups.
